# Assessing the Accessibility of Medical Facilities with Multilingual Support for Foreign Residents in Japan: A Spatial Distribution Perspective

**DOI:** 10.1101/2025.03.24.25324566

**Authors:** Yuchen Lu

## Abstract

In recent years, as the population of foreign residents in Japan has rapidly increased, societal discussion about their medical coverage has intensified. Given the concerning health status and medical service utilization of foreign residents in Japan, and the critical role of medical facilities with multilingual support (MFMS)—including key facilities designated for foreign patients—this study is the first to assess the accessibility of such facilities for foreign residents at the municipal level using publicly available geographic information systems. The evaluation focused on two departments closely tied to daily life: surgery and internal medicine. The results reveal that accessibility indices for medical facilities offering multilingual services in surgery are low across most areas of Hokkaido and the Tohoku region, as well as northern Niigata and Shizuoka prefectures, central Miyazaki prefecture, and major islands with foreign resident populations. Similar patterns were observed for internal medicine. This suggests that local foreign residents may face significant challenges in accessing specialized, linguistically consistent medical services in surgery or internal medicine.

## Introduction

It is widely recognized that Japan faces significant demographic challenges, including an aging population and declining birthrates. In recent years, the Japanese government has prioritized recruiting foreign labor as a key strategy for national development to address the severe domestic labor shortage. This approach involves introducing new labor visa categories and relaxing residency requirements for work visas. By the end of 2023, the number of foreign residents in Japan reached an all-time high of 3.41 million. As this population grows rapidly, societal concern for their social security, particularly medical care, has intensified. Research shows that, even when facing serious health issues, these residents often avoid local medical services, preferring self-treatment or enduring pain [1, 2]. Japan’s healthcare system currently fails to adequately safeguard their health. Without reforms to the medical system, disparities in healthcare access and health outcomes may widen further [3].

The 2020 global Coronavirus Disease 2019 (COVID-19) pandemic is considered to have further constrained access to healthcare for foreign residents or immigrants, disproportionately affecting their health. Due to poorer living and working conditions, immigrants face a higher risk of contracting COVID-19 and a greater likelihood of severe outcomes [4]. During the pandemic, it was found that immigrant students were more prone to depression and other mental health issues compared to non-immigrant students [5]. Some scholars advocate using this period as an opportunity to implement regular, extensive health surveys among immigrants [6]. From a social equity perspective, there is a gradually increasing international focus on the health rights of foreign residents.

The poor health of foreign residents and their difficulty in accessing high-quality medical services can result in internalities and negative externalities. Significant health disparities between different social groups often lead to disadvantaged groups lacking the necessary resources for social and economic participation [7]. Furthermore, foreign residents’ limited cultural and linguistic understanding of local COVID-19 prevention and treatment information may elevate their health risks, which are likely to impact native populations as well [8]. Some studies argue that providing high-quality medical services to foreign residents not only facilitates their active integration into local society but also boosts their productivity [9].

Language barriers are widely acknowledged as a major external factor that impedes foreign residents’ access to high-quality healthcare [10]. These barriers significantly hinder the effective communication of information [11]. Research indicates that patients facing language obstacles tend to seek medical care more frequently, which increases treatment costs and reduces the efficiency of healthcare workers [12]. Therefore, addressing or mitigating language barriers is essential in efforts to reduce health disparities. Providing language-concordant care is considered an effective method to significantly improve communication between healthcare providers and patients, as well as to enhance patient satisfaction [13].

Currently, to promote the elimination of language barriers and the widespread adoption of language-concordant care, Japanese prefectural governments have selected certain medical institutions from among those willing to assist foreign patients. These institutions have been designated as key medical facilities for receiving foreign patients. Key medical facilities typically receive government funding to improve the quality of multilingual medical services by establishing international patient departments, training international patient care coordinators, and enhancing medical translation systems.

Additionally, these key facilities may also assist in fulfilling the demands for multilingual medical services raised by neighboring medical institutions [14]. This demonstrates that key medical facilities for receiving foreign patients play a crucial role in integrating medical interpreting resources and efficiently providing language-concordant care to foreign patients.

In addition to key medical facilities, two non-profit organizations—Medical Excellence Japan (MEJ) and the Japan Medical Education Foundation (JMEF)—have established certification systems for large medical institutions that were among the first to systematically receive and treat international patients under an initiative to globalize Japanese healthcare services. MEJ evaluates medical institutions to assess both their readiness to receive international patients and their experience in providing such services. Institutions that meet these standards are designated as Japan International Hospitals (JIH). Similarly, JMEF has established the Japan Medical International Patient Service Certification (JMIP), aimed at certifying medical institutions capable of providing multilingual diagnostic and treatment services to meet the needs of foreign patients.

Figure 1 presents results from a Ministry of Health, Labour and Welfare of Japan (MHLWJ) survey conducted in September 2022, targeting hospitals and clinics nationwide regarding their acceptance of foreign patients. The findings show that the proportion of key medical facilities for receiving foreign patients and those accredited with JMIP or JIH is significantly higher than the national average. In this study, we refer to these three types of facilities—each possessing professional qualifications in providing multilingual medical services—as medical facilities with multilingual support (MFMS)^1^. Given that patients in Japan have considerable autonomy in selecting healthcare providers [15], Figure 1 suggests that foreign patients in Japan tend to prefer MFMS for their healthcare needs.

**Fig 1.**
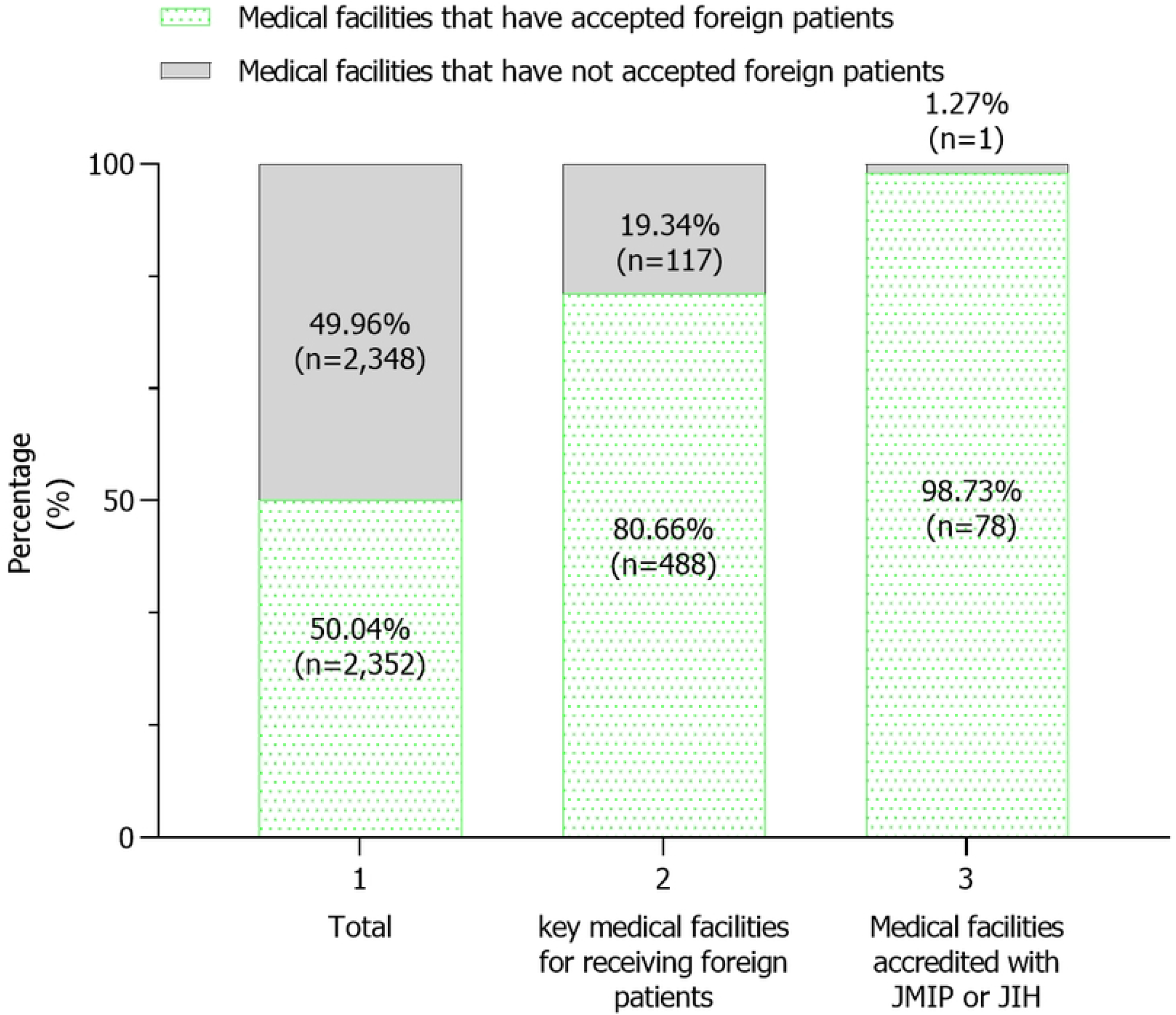
Number of medical facilities that have accepted foreign patients. Made by the author based on the “Survey on the Acceptance of Foreign Patients in Medical Facilities for Reiwa 4 years (Summary Version)”

Based on the discussion above, it is evident that MFMS play a crucial role in providing medical coverage for foreign residents in Japan, and researching the accessibility of these facilities is of significant practical importance for improving the health status of this group. Currently, research on the medical accessibility of foreign residents in Japan primarily focuses on social organizational factors, such as the socio-economic characteristics that hinder this group’s use of medical services. However, research on medical accessibility must also account for geographical factors, including the travel time and distance required to access healthcare [16]. Despite this need, there remains limited research on the spatial accessibility of these facilities for foreign residents in Japan.

With the disclosure of the list of key medical facilities for foreign patients by the end of 2023, exploring the spatial accessibility of MFMS for foreign residents has become feasible. Considering the anticipated growth and permanent residency trends among foreign populations in Japan, and the accompanying increase in medical needs, this study aims to examine the spatial distribution and accessibility of MFMS that play a crucial role in healthcare for these populations. It will focus on multilingual medical services in surgery and internal medicine—areas of potentially high demand and closely related to everyday life. The findings will offer policy recommendations for the regional planning of medical facilities to ensure more effective allocation.

## Data and Methods

### Data

To facilitate easy access to medical services in Japan for visitors and foreign residents, “A Compiled List of Information on Medical Facilities That Accept Foreign Patients” (hereinafter referred to as “the list”) was released in December 2023. This list provides detailed information on key medical facilities dedicated to serving foreign patients. It includes, but is not limited to, geographical details (postal codes and addresses) of each facility, clinical departments equipped to treat foreign patients, the type of facility (hospital or clinic), the presence of international patient departments, the availability of international patient care coordinators and medical interpreters, and support for remote medical interpretation services. Regarding medical facilities accredited with JMIP, as of December 2023, there are a total of 61 such institutions, all of which are general hospitals. Each hospital incorporates an international patient department, ensuring that foreign patients can access a range of clinical departments without barriers. Information on these hospitals, including their geographical locations, is publicly disclosed on the JMEF official website. On the other hand, as of December 2023, there are 43 medical facilities accredited with JIH, which are also general hospitals equipped with international patient departments. The MEJ official website hosts a dedicated search page for these medical facilities, providing comprehensive details, including location information.

Regarding data on the foreign resident population, this study utilizes a database named “Statistics on Foreign National Residents”, periodically released by the Immigration Services Agency of Japan. This database is updated biannually, and our analysis will be based on the latest data published in July 2024, which reflects the status as of December 2023. This semiannual data encompasses several segmented subsets, among which the statistics of foreign residents’ population by municipalities offer more granular administrative unit-level population data. Additionally, as the basic bureaucratic administrative divisions, various types of data can be correlated and matched at the municipality level^2^. Further information regarding the dataset used in this study can be found in Table 1.

**Table 1.**
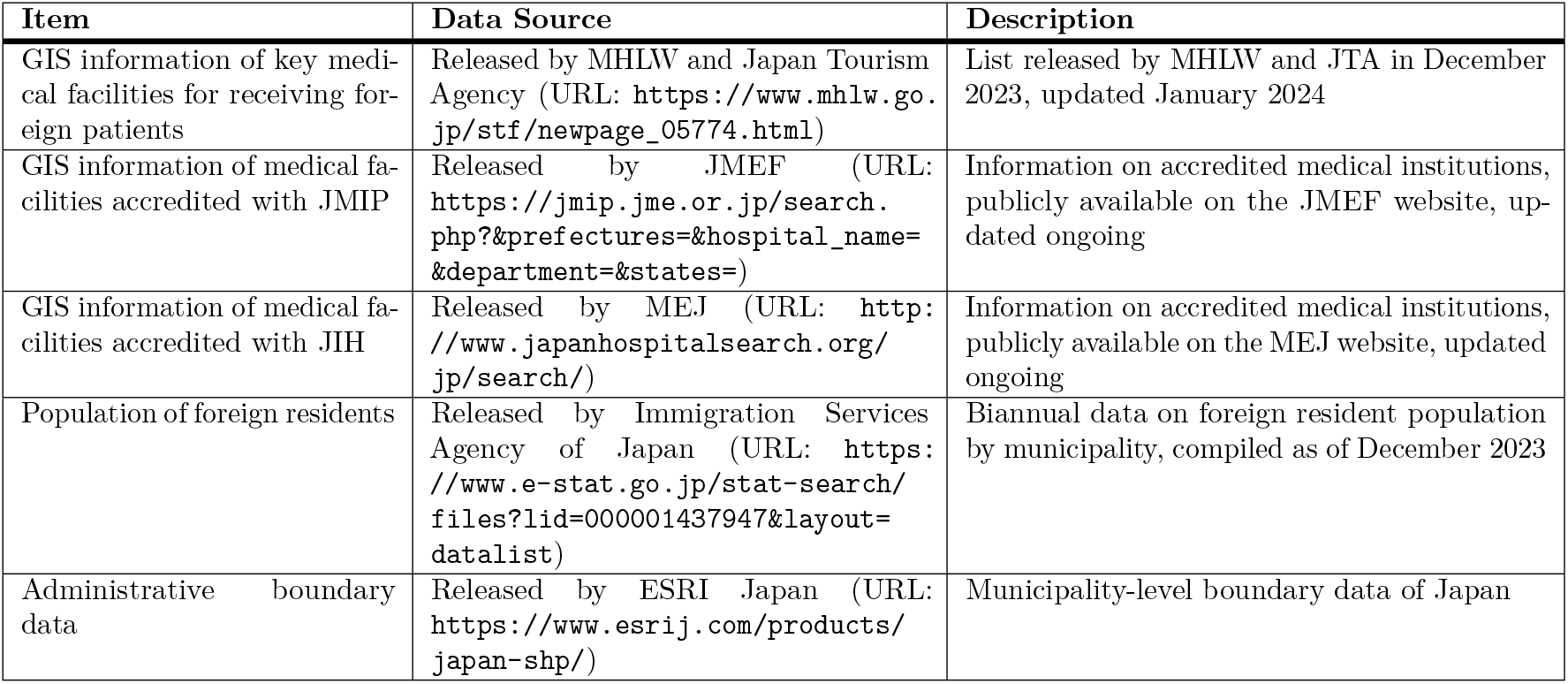
Dataset Summary for Spatial Accessibility Analysis of Multilingual Healthcare Institutions in Japan.

In the list, we will use the information in the column “Departments that can accommodate foreign patients” to filter the target MFMS. This includes institutions explicitly marked as providing surgical or internal medicine services in that column, as well as those marked as offering all diagnostic and treatment services. Additionally, for medical facilities accredited with JMIP or JIH, we will use the department search system on the official websites to identify institutions that provide general internal medicine or general surgery services. It should be noted that some institutions may appear on the list and also have JMIP or JIH accreditation, sometimes even holding dual certifications. In such cases, since the list provides more detailed information on multilingual medical service departments, we will rely on the list to determine whether the institution is a target facility.

## Methods

First, this study utilizes GIS software called ArcGIS Pro to map the spatial distribution of MFMS and foreign residents in Japan, establishing a foundation for subsequent analysis of spatial accessibility. It is crucial to note that the addresses of MFMS, specifically those accredited with JMIP or JIH, must be individually collected from their official websites. Similar to key medical facilities for receiving foreign patients, these institutions’ addresses are typically presented in the Japanese address format. Therefore, these addresses must first be converted into geographic coordinates (latitude and longitude) through geocoding. For the geocoding process, this study leverages a free online platform provided by the Center for Spatial Information Science, The University of Tokyo, called CSV Geocoding Service^3^. It should be noted that, during the geocoding process, the precision of the longitude and latitude coordinates obtained for a small number of medical facilities may be compromised due to the excessive length of their Japanese address format, thereby preventing the achievement of the highest precision^4^. In response to this issue, this study utilized a manual approach, with the use of Google Maps to obtain the geographic coordinates^5^.

Given that the target population of this study is the potential demand group, specifically all foreign residents in Japan, the assessed spatial accessibility is best classified as potential spatial accessibility. The measurement of this accessibility has traditionally relied on the gravity model [17], which quantifies accessibility based on service capacity and the distance or travel time between locations (1).

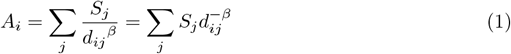

However, this model mainly considers supply factors and overlooks the demand side. To address this limitation, the gravity model was modified by incorporating population data at each location [18], enhancing its theoretical accuracy while increasing computational complexity. This modification increases the complexity, particularly when adjusting the distance friction coefficient *β*, which varies according to the development level of regional transportation networks. In response to these complexities, the Floating Catchment Area (FCA) method was developed as a simplified alternative that omits *β* and assumes uniform access within a given radius. However, the FCA method has its drawbacks: it assumes that all potential users within a circular catchment area, centered at location *i* and with a radius equivalent to the distance threshold *d*, have an equal probability of accessing services at a service point. This assumption is flawed, as some users within the catchment area might be located farther from the service point, beyond the threshold distance, meaning they cannot access the services. Moreover, service points near the periphery of the catchment area might preferentially serve users outside the catchment area who are closer in distance or travel time than those within the area [19].

Service point geography was optimized to enhance service equity by creating catchment areas with equal service probabilities [20]. In overlapping areas, the service probability for intersecting sections was proposed as the arithmetic sum of overlapping probabilities [20], refining the FCA method’s assumptions and improving probability calculations. Building on these insights, the FCA method was enhanced, leading to the development of the two-step floating catchment area (2SFCA) method [19]. The 2SFCA method is described as consisting of two primary steps [21]. **Step 1** involves searching for all potential patients at location *k* within a threshold travel time or distance *d*_0_ for each healthcare provider’s location *j*, and calculating the ratio of healthcare providers to potential patients *R*_*j*_ within that catchment area (2).

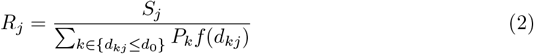

The function *f* (*d*_*kj*_) in the denominator is referred to as the distance decay function, which represents the variation in the probability of physicians or medical facilities accessing potential patients as travel time changes. **Step 2** entails identifying all healthcare providers at location *k* within the threshold travel time or distance *d*_0_ for each potential patient’s location *i*, and then summing the ratios of healthcare providers to potential healthcare service users calculated in the **Step 1** (3).

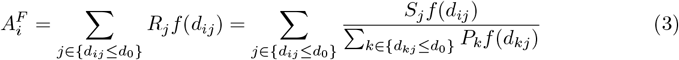

The index 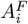 represents the potential spatial accessibility of the population at location *i* to healthcare providers, as measured by the 2SFCA method. The distance decay function *f* (*d*_*ij*_) denotes the function that represents the change in the probability of potential patients reaching physicians or medical facilities as the travel time varies. Both distance decay functions *f* (*d*_*ij*_) and *f* (*d*_*kj*_) are considered to diminish with an increase in travel time or distance [22]. When *f* (*d*_*ij*_) = *f* (*d*_*kj*_) = 1, the calculations for Equations (2) and (3) are substantially simplified, and under these conditions, the 2SFCA method is also referred to as the original 2SFCA method (4).

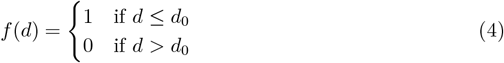

Compared to traditional place-based accessibility measures, the original 2SFCA method more comprehensively accounts for the interactions between supply and demand. Importantly, this method is not constrained by the friction-of-distance coefficient [23]. Due to these advantages, it is extensively utilized to evaluate spatial distribution inequalities among healthcare service providers or medical institutions [24, 25].

Regarding the *f* (*d*), beyond the dichotomous form, commonly utilized function types include the Power function, Gaussian function, and Kernel density function [26]. The determination of *f* (*d*) and *d*_0_ has been thoroughly discussed [23]. Research shows that the coefficient of variation *C*_*v*_ of different *f* (*d*) manifestations fluctuates with changes in *d*_0_, but begins to converge around *d*_0_ = 10 miles (approximately 16 kilometers), stabilizing as *d*_0_ increases further. Additionally, it is noted that for larger-scale study areas, setting *d*_0_ above a certain threshold is essential to ensure consistent spatial patterns [23].

In determining *d*_0_ for this study, we relied on findings from the Survey on No-doctor Districts conducted in Heisei 26 (2014). This survey showed that in regions with limited medical facilities, the average travel distance by car to the nearest medical institution is 23.3 kilometers. Drawing on prior research and considering the study area’s scope, we adopted the original 2SFCA method, using a binary distance decay function with a 25-kilometer threshold to evaluate the spatial accessibility of MFMS in internal medicine and surgery. Additionally, due to unavailable data on the number of physicians providing multilingual services in surgery or internal medicine at each facility, we assigned a uniform provider capacity of 1.

## Results

Fig 2 shows the geographic distribution of foreign residents across various municipalities in Japan as of December 2023. The figure clearly indicates that foreign residents in Japan are primarily concentrated in economically developed areas such as Tokyo, Osaka, and Aichi prefectures. However, apart from a few areas where foreign residents are not recorded, the majority of municipalities have foreign resident populations. This suggests that while the geographic distribution of foreign residents is partially concentrated in specific areas, it is also widely dispersed, exhibiting an uneven distribution pattern.

**Fig 2.**
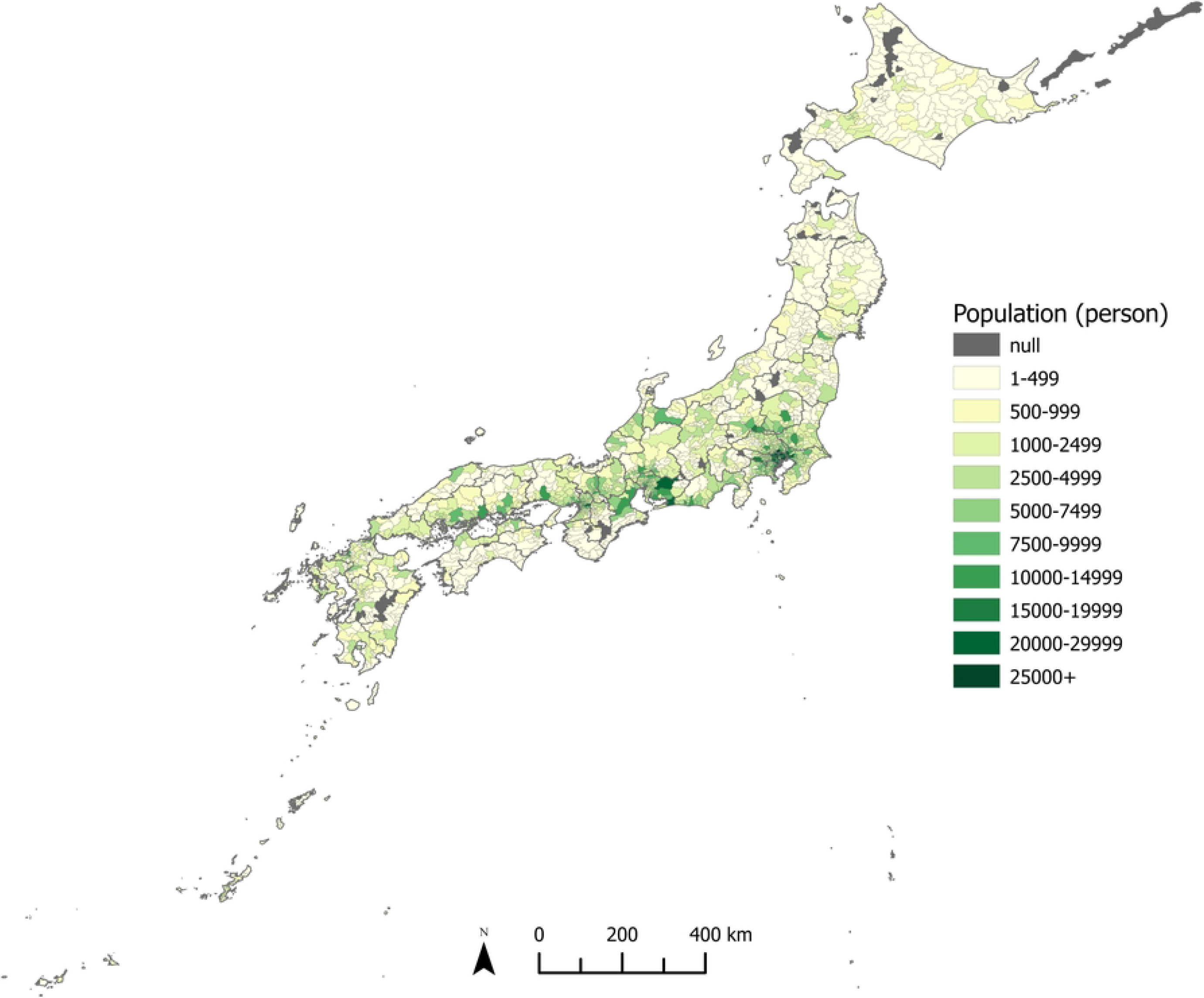
Distribution of Foreign Residents by Municipality in Japan, End of 2023. Made by the author using ArcGIS Pro 3.2.1.

Figs 3 and 4 illustrate the geographic distribution of MFMS offering multilingual medical services in surgery and internal medicine, respectively. Nationwide, 861 MFMS provide these services in surgery, and 1,332 do so in internal medicine. The distribution of MFMS closely aligns with that of foreign residents, concentrating primarily in densely populated and economically developed areas where foreign populations also cluster.

**Fig 3.**
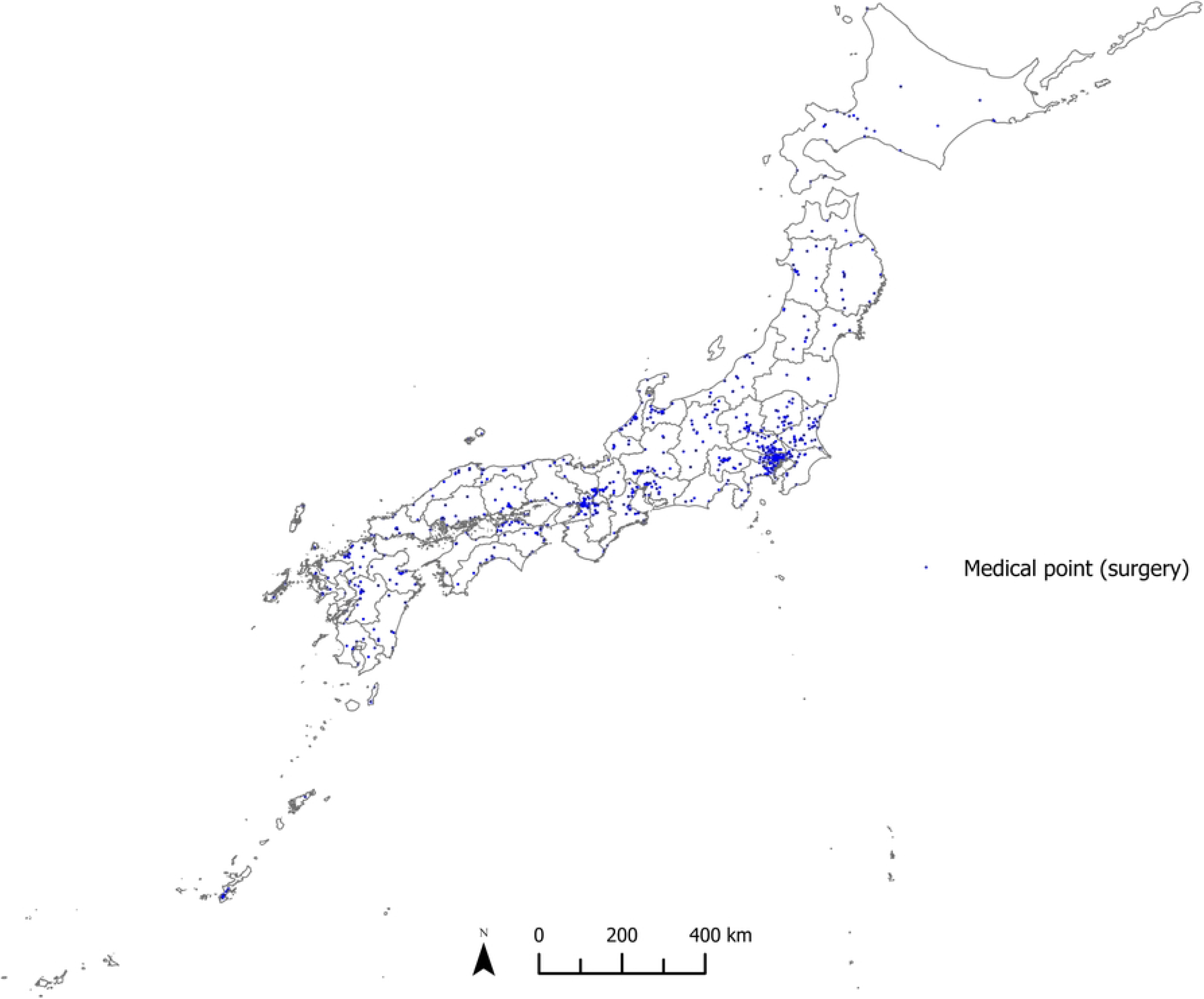
Geographical Distribution of MFMS for Surgical Departments. Made by the author using ArcGIS Pro 3.2.1.

**Fig 4.**
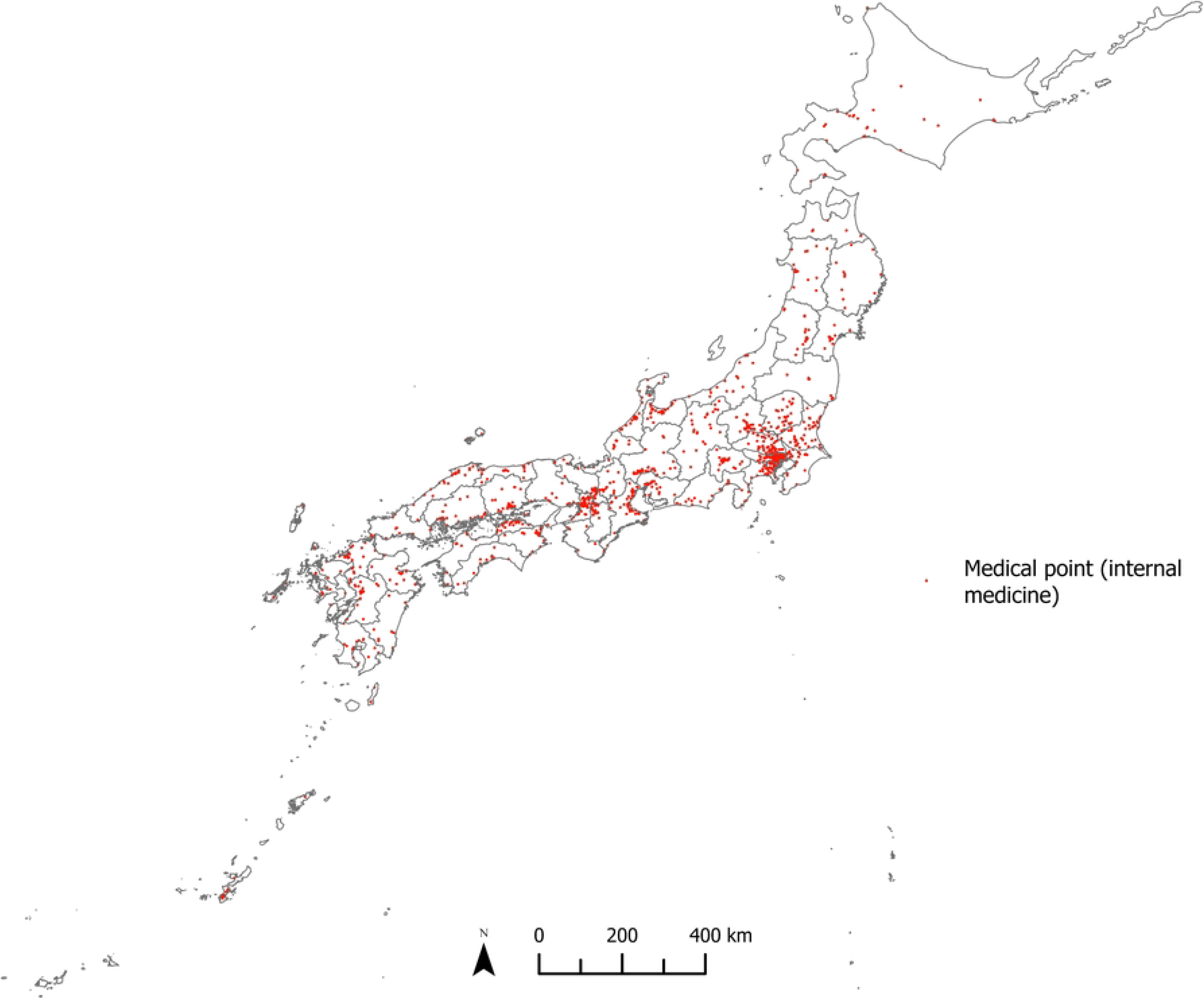
Geographical Distribution of MFMS for Internal Medicine Departments. Made by the author using ArcGIS Pro 3.2.1.

However, regional distribution exhibits significant heterogeneity. Among MFMS offering multilingual services in surgery, Tokyo has 87, while Miyagi and Saga prefectures have only 4 each. Similarly, the disparity is even more pronounced for MFMS offering these services in internal medicine: Tokyo has 205, whereas Aomori and Saga prefectures have just 4 each.

Figs 5 and 6 display the spatial distribution of MFMS accessibility scores under a 25-kilometer mobility distance threshold. These figures highlight significant heterogeneity in MFMS accessibility across regions. Areas colored white indicate an accessibility score of 0, denoting poor accessibility; darker colors represent higher accessibility. For MFMS offering multilingual services in surgery, accessibility scores of 0 are observed across extensive areas of Hokkaido, northern Aomori, central Iwate, western Yamagata, most of Fukushima, northern Niigata, northern Shizuoka, central Miyazaki, and several remote islands, reflecting limited access to surgical multilingual services for foreign residents in these regions. The pattern for internal medicine mirrors that of surgery, with scores of 0 in vast areas of Hokkaido, parts of the Tohoku region, northern Shizuoka, central Miyazaki, and several remote islands. Conversely, while areas with high foreign resident concentrations generally lie within the range of multilingual medical services, the accessibility scores in densely populated regions such as Tokyo, Osaka, and Aichi prefectures remain notably low.

**Fig 5.**
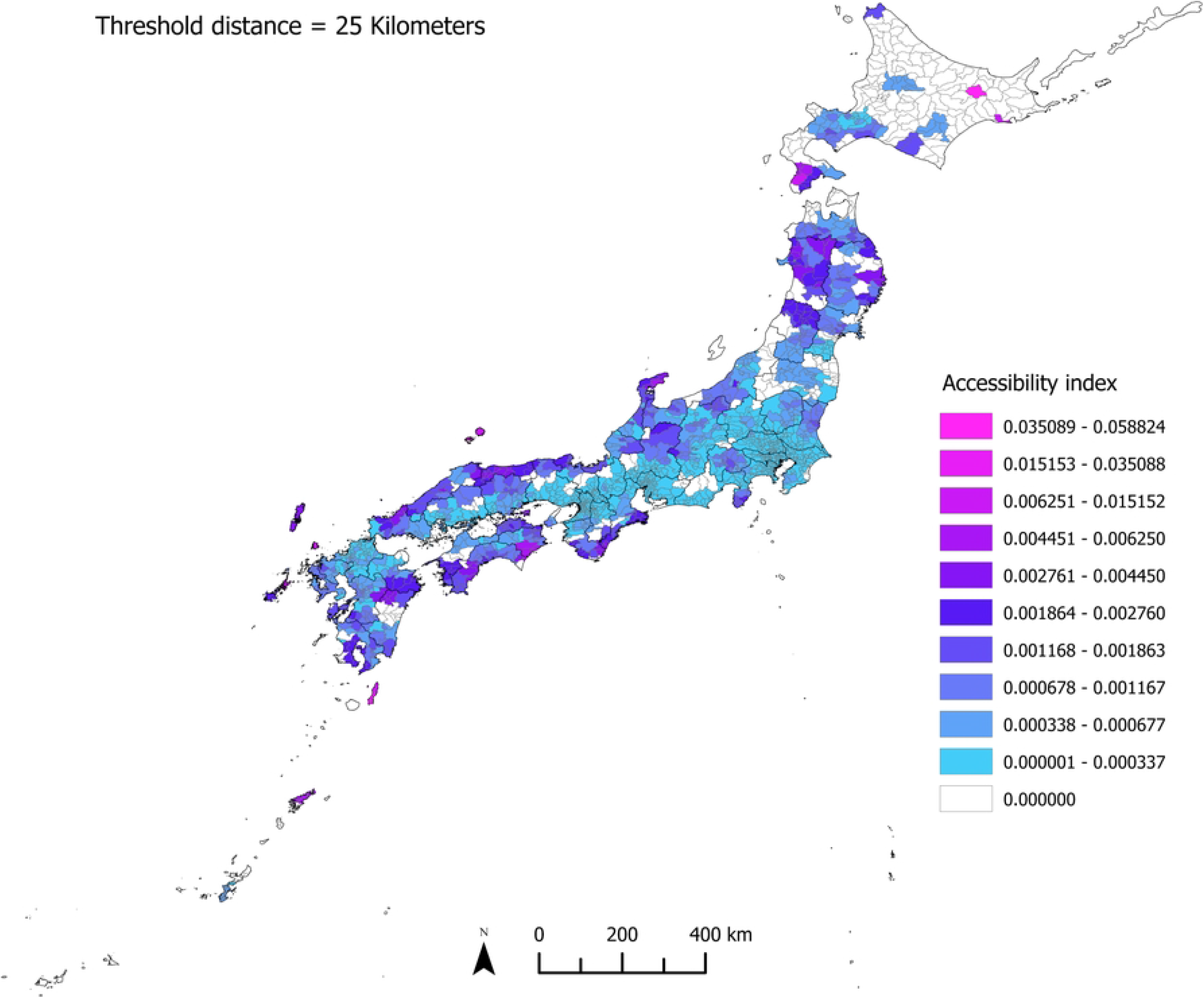
Spatial Distribution of Accessibility Indices for Surgery. Made by the author using ArcGIS Pro 3.2.1.

**Fig 6.**
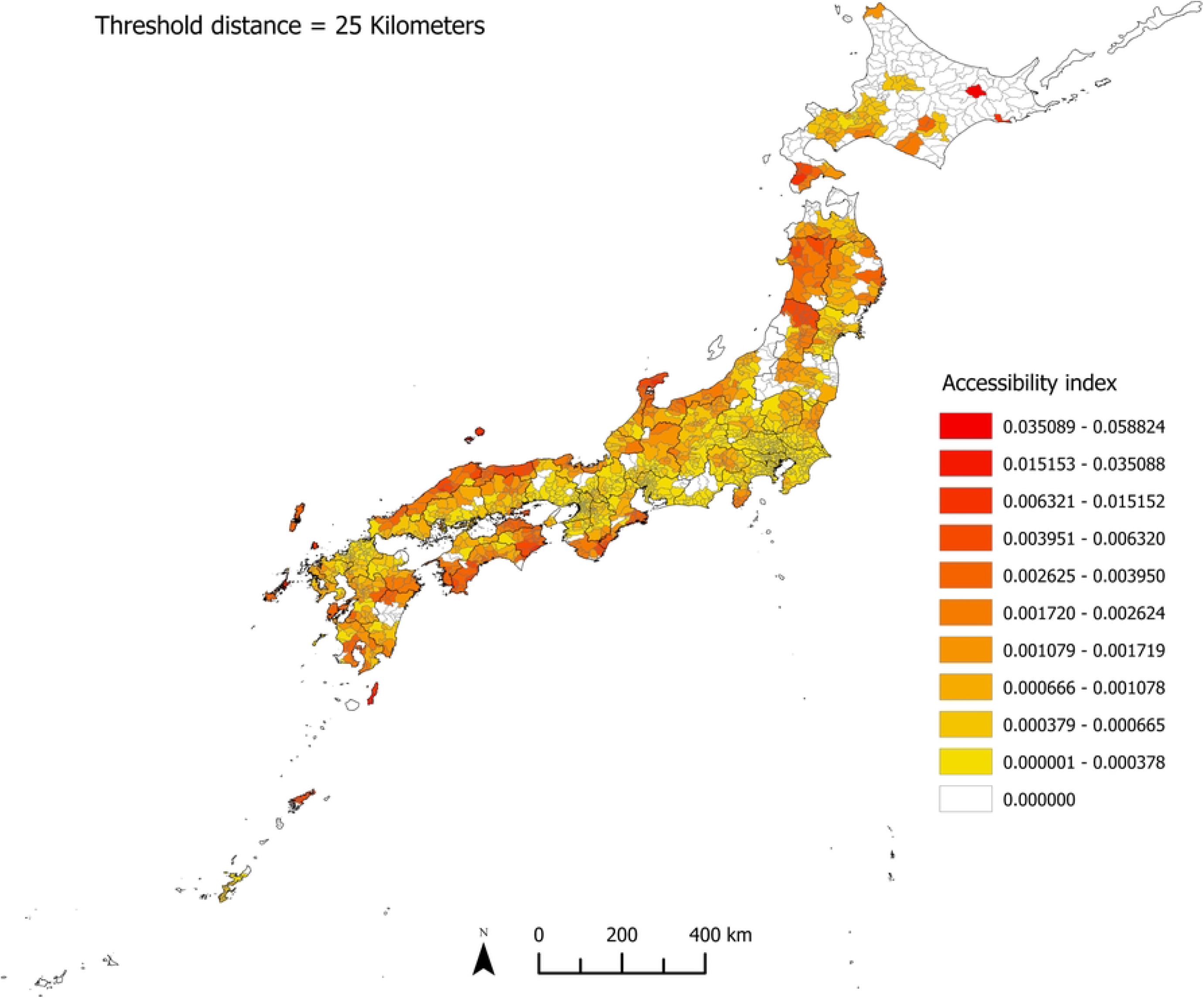
Spatial Distribution of Accessibility Indices for Internal Medicine. Made by the author using ArcGIS Pro 3.2.1.

## Discussion

We should focus on areas with lower accessibility. Overall, most areas of Hokkaido, parts of the Tohoku region, northern Niigata and Shizuoka prefectures, central Miyazaki, and some remote islands have low accessibility scores. This suggests that foreign residents in these areas are less likely to access highly specialized, linguistically consistent medical services in surgery or internal medicine. To obtain such services in specific departments, local foreign residents may need to travel longer distances, increasing their economic burden and time costs. Therefore, in areas with relatively low medical resource density—such as Hokkaido, the Tohoku region, and Niigata—it is especially important to equitably distribute multilingual medical service resources to improve coverage. For example, in the Okhotsk region of Hokkaido, even with a travel distance threshold of 25 kilometers, the accessibility index remains zero for most municipalities, indicating a near absence of MFMS in this area and its vicinity. However, the number of hospital beds and doctors per 100,000 people in this region does not lag significantly behind other parts of Hokkaido [27]. As for lower accessibility scores in densely populated areas, this may result from the high number of foreign residents and greater potential medical demand, rendering the available medical facilities relatively insufficient.

Given that most MFMS are designated by prefectural governments as key facilities for receiving foreign patients, policymakers should prioritize the rational spatial layout and balanced allocation of resources when advancing multilingual medical services.

Regarding the limitations of this study’s methods and processes, it should first be noted that while most MFMS are designated as key facilities for foreign patients by prefectural governments, some facilities capable of providing multilingual services are excluded from the list for various reasons. This lack of data may introduce bias into the estimated accessibility indices. A second limitation arises from the inability to obtain age distribution data for foreign residents across municipalities; this study thus uses the total foreign resident population in each area as the potential demand population.

However, certain age groups, such as children under 15, may require pediatric care rather than surgical or internal medicine services when ill, potentially underestimating the actual demand population. For future improvements, as mentioned earlier, medical facilities offering multilingual support could enhance services through means beyond staff language training, such as introducing translation devices. In this study, we simplified the analysis by not considering whether the language services provided by medical institutions match the native languages of the potential population. In future research, we plan to incorporate data on available translation languages at medical institutions and the nationalities of foreign residents to refine the analysis further.

## Conclusion

This study is the first to explore the spatial distribution and accessibility of medical facilities in Japan that offer multilingual medical services in surgery and internal medicine. The findings reveal an imbalance in the spatial distribution of such medical facilities, indicating that foreign residents in certain areas may face significant barriers in accessing these services.

## Data Availability

All relevant data are within the manuscript and its Supporting Information files.

## Footnotes

1 Note that a medical facility may simultaneously serve as a key medical facility for receiving foreign patients and hold at least one accreditation from JMIP or JIH.

2 Japan’s administrative system comprises a three-tier structure. At the apex is the national government, succeeded by the prefectural governments. The third tier of administration is comprised of municipalities. Therefore, municipalities are the basic units of administrative divisions in Japan, numbering approximately 1,700.

3 CSV Geocoding Service usage agreement (Accessed on November 20, 2023) URL: https://geocode.csis.utokyo.ac.jp/home/csv-admatch/

4 The CSV Geocoding Service provides eight different precision levels ranging from 0 to 7 for the converted longitude and latitude, with level 7 representing the highest precision.

5 Google Map Usage Agreement (Accessed on February 11, 2024) URL: https://about.google/brand-resourcecenter/products-andservices/geo-guidelines/

## Notes

### Competing Interest Statement

The authors have declared no competing interest.

### Funding Statement

The author(s) received no specific funding for this work.

### Author Declarations

This study does not involve human or animal testing, nor does it use data derived from human subjects or identifiable private information.

## References

1. Kunii, O, Nomiyama, K. Present Status of Medical Care for Foreigners in Tochigi Prefecture, Japan (2) Illness Behavior of Foreign Workers. Nippon Eiseigaku Zasshi (Japanese Journal of Hygiene). 1993;48(3):685–691. (Japanese)

2. Morita, N, Kanamori, M, Nochi, M, Kondo, N. A mixed methods study on specifying the inhibitory factors to access medical services and effective support for foreign residents living in Japan. Kokusai Hoken Iryo (Journal of International Health). 2021; 36(3):107–121. (Japanese)

3. Yasukawa, K, Sawada, T, Hashimoto, H, Jimba, M. Health-care disparities for foreign residents in Japan. The Lancet. 2019;393(10174):873–874.

4. OECD. What has been the impact of the COVID-19 pandemic on immigrants? An update on recent evidence. OECD Policy Responses to Coronavirus (COVID-19), OECD Publishing, Paris.

5. Pieh, C, Dale, R, Jesser, A, Probst, T, Plener, P, Humer, E. The impact of migration status on adolescents’ mental health during COVID-19. In Healthcare. 2022;10(176):176.

6. Lebano, A, Hamed, S, Bradby, H, Gil-Salmerón, A, Durá-Ferrandis, E, Garcés-Ferrer, J, Azzedine, F, Riza, E, Karnaki, P, Zota, D. Migrants’ and refugees’ health status and healthcare in Europe: a scoping literature review. BMC Public Health. 2020;20:1–22.

7. Woodward, A, Kawachi, I. Why reduce health inequalities? Journal of Epidemiology and Community Health. 2000;54(12):923–929.

8. Kluge, H, Jakab, Z, Bartovic, J, d’Anna, V, Severoni, S. Refugee and migrant health in the COVID-19 response. The Lancet. 2020;395(10232):1237–1239.

9. Lebrun, L. Effects of length of stay and language proficiency on health care experiences among immigrants in Canada and the United States. Social Science & Medicine. 2012;74(7):1062–1072.

10. Teraoka, M, Muranaka, Y. Aspects of cross-cultural experience perceived by foreigners living in Japan when using its healthcare services. Journal of Japan Academy of Nursing Science. 2017;37:35–44. (Japanese)

11. Kim, G, Loi, C, Chiriboga, D, Jang, Y, Parmelee, P, Allen, R. Limited English proficiency as a barrier to mental health service use: A study of Latino and Asian immigrants with psychiatric disorders. Journal of Psychiatric Research. 2011;45(1):104–110.

12. Scheppers, E, Van Dongen, E, Dekker, J, Geertzen, J, Dekker, J. Potential barriers to the use of health services among ethnic minorities: a review. Family Practice. 2006;23(3):325–348.

13. Detz, A, Mangione, C, de Jaimes, F, Noguera, C, Morales, L, Tseng, C, Moreno, G. Language concordance, interpersonal care, and diabetes self-care in rural Latino patients. Journal of General Internal Medicine. 2014;29:1650–1656.

14. Nicola, F, Nishimura, A. Progress and challenges in medical interpreting services in Japan. Migration Policy Review. 2016;8:193–203. (Japanese)

15. Teruyuki, K. Japan’s healthcare delivery system: From its historical evolution to the challenges of a super-aged society. Global Health & Medicine. 2024;6(1):6–12.

16. Khan, AA. An integrated approach to measuring potential spatial access to health care services. Socio-Economic Planning Sciences. 1992;26(4):275–287.

17. Hansen, W. How accessibility shapes land use. Journal of the American Institute of Planners. 1959;25(2):73–76.

18. Joseph, A, Bantock, P. Measuring potential physical accessibility to general practitioners in rural areas: A method and case study. Social Science & Medicine. 1982;16(1):85–90.

19. Luo, W, Wang, F. Measures of spatial accessibility to health care in a GIS environment: synthesis and a case study in the Chicago region. Environment and Planning B: Planning and Design. 2003;30(6):865–884.

20. Radke, J, Mu, L. Spatial decompositions, modeling and mapping service regions to predict access to social programs. Geographic Information Sciences. 2000;6(2):105–112.

21. Luo, W, Whippo, T. Variable catchment sizes for the two-step floating catchment area (2SFCA) method. Health & Place. 2012;18(4):789–795.

22. Wang, F, Minor, W. Where the jobs are: Employment access and crime patterns in Cleveland. Annals of the Association of American Geographers. 2002;92(3):435–450.

23. Chen, X, Jia, P. A comparative analysis of accessibility measures by the two-step floating catchment area (2SFCA) method. International Journal of Geographical Information Science. 2019;33(9):1739–1758.

24. Kanuganti, S, Sarkar, A, Singh, A. Quantifying accessibility to health care using two-step floating catchment area method (2SFCA): A case study in Rajasthan. Transportation Research Procedia. 2016;17:391–399.

25. Khashoggi, B, Murad, A. Use of 2SFCA method to identify and analyze spatial access disparities to healthcare in Jeddah, Saudi Arabia. Applied Sciences. 2021;11(20):9537.

26. Wang, F, Wang, K. Measuring spatial accessibility to ecological recreation spaces in the Pearl River Delta region: an improved two-step floating catchment area method. Journal of Spatial Science. 2018;63(2):279–295.

27. Akino, T. Medical care in local community of Hokkaido in the drastic change of medical treatment system. Nippon Ronen Igakkai Zasshi. Japanese Journal of Geriatrics. 2007;44(5):556–563. (Japanese)

